# Evaluation of *in silico* pathogenicity prediction tools for the classification of small in-frame indels

**DOI:** 10.1101/2022.10.27.22281598

**Authors:** S. Cannon, M. Williams, A. C. Gunning, C. F. Wright

## Abstract

**Background:** The use of *in silico* pathogenicity predictions as evidence when interpreting genetic variants is widely accepted as part of standard variant classification guidelines. Although numerous algorithms have been developed and evaluated for classifying missense variants, in-frame insertions/deletions (indels) have been much less well studied.

**Methods:** We created a dataset of 3964 small (<100bp) indels predicted to result in in-frame amino acid insertions or deletions using data from gnomAD v3.1 (minor allele frequency of 1-5%), ClinVar and the Deciphering Developmental Disorders (DDD) study. We used this dataset to evaluate the performance of nine pathogenicity predictor tools: CADD, CAPICE, FATHMM-indel, MutPred-Indel, MutationTaster2 PROVEAN, SIFT-indel, VEST-indel and VVP.

**Results:** Our dataset consisted of 2224 benign/likely benign and 1740 pathogenic/likely pathogenic variants from gnomAD (n=809), ClinVar (n=2882) and, DDD (n=273). We were able to generate scores across all tools for 91% of the variants, with areas under the ROC curve (AUC) of 0.81-0.96 based on the published recommended thresholds. To avoid biases caused by inclusion of our dataset in the tools’ training data, we also evaluated just DDD variants not present in either gnomAD or ClinVar (70 pathogenic and 81 benign). Using this subset, the AUC of all tools decreased substantially to 0.64-0.87. Overall, VEST-indel performed best, with AUCs of 0.93 (full dataset) and 0.87 (DDD subset).

**Conclusions:** Algorithms designed for predicting the pathogenicity of in-frame indels perform well enough to aid clinical variant classification in a similar manner to missense prediction tools.

## BACKGROUND

Next generation DNA sequencing (NGS) is transforming healthcare by facilitating novel understanding of disease and uptake of precision medicine initiatives (1, 2). Genetic variation is widespread, with every individual carrying >200 very rare coding variants (3), so molecular diagnosis of monogenic disorders requires expert clinical and scientific interpretation of variants detected by NGS. Classifying the pathogenicity of candidate causal variants is essential for robust diagnosis and management of genetic disorders. To this end, numerous *in silico* pathogenicity prediction algorithms have been developed and are widely used as evidence when interpreting genetic variants. The use of pathogenicity predictors is supported by current guidelines from the American College of Medical Genetics and Genomics (ACMG) and Association for Molecular Pathology (AMP) (4) and, more recently, the UK Association for Clinical Genomic Science (ACGS) **(5)**, through the PP3/BP4 criteria. Pathogenicity prediction algorithms incorporate various lines of evidence to predict the impact of variation on protein function, including evolutionary inter-species sequence conservation (6), physico-chemical distances between amino acids (7) as well as integrated tests for identifying regulatory features (8). Some also incorporate human variation and disease data (9) by querying gene or variant level understanding from open source (10) or proprietary (11) databases. These aggregated data are used to generate statistical prediction models, such as supervised machine learning classifiers (12), which produce a score used to assign pathogenicity status to a given variant.

Most pathogenicity predictors have been developed to predict the effect of missense substitutions (13, 14), which are primarily caused by single nucleotide variants (SNVs) in the protein-coding regions of the genome. However, small insertions and deletions (indels) account for between 13-18% of all variation in the human genome (15, 16), both within and outside protein-coding regions, and have been linked to numerous rare heritable diseases (17) as well as cancerous somatic mutations(18). Approximately 40% of coding indels are in-frame (19), defined as a nucleotide length (n), wholly divisible by three, which results in the removal or addition of n/3 amino acids. Unlike frame-shifting indels, which are generally assumed to cause loss-of-function, the insertion or deletion of a small number of amino acids is likely to have a similarly deleterious effect on a protein as substitution of one amino acid for another. Indeed, missense variants and in-frame indels are frequently grouped together as “protein altering variants” and overall assumed to have “moderate” impact (20).

Numerous small in-frame indels have been shown to cause monogenic disease, most famously (p.Phe508del) in *CFTR* (21). However, in general, the classification of in-frame indels has been much less well studied than missense and loss-of-function variants. To this end, we created a novel dataset of previously classified in-frame indels, constructed from three databases, two open source (gnomAD (22) and, ClinVar (10)) and one managed access (Diagnosing Developmental Disorders study (DDD) (23)), and use this dataset to evaluate the performance of nine *in silico* prediction algorithms. We show that although the accuracy of pathogenicity classifiers varies across tools, overall the performance is comparable to those designed for missense variants.

## METHODS

### Benchmark dataset generation

Variants were retrieved from ClinVar (10), the DDD study (23), and gnomAD (v3.1.1) (22) (accessed 18/03/2021) before filtering for suitability for this study (**Figure 1**). Briefly, variants in genome build GRCh38 were included if they were evenly divisible by 3 and <100 base-pairs in length. Assumed benign variants with a minor allele frequency 1-5% were retained from the gnomAD population database, while variants classified as likely pathogenic (LP), pathogenic (P), benign (B) or likely benign (LB) were retained from the two clinical datasets. Identical variants in more than one database were retained from only one using the preferential order of DDD, ClinVar then gnomAD, and variants with conflicting annotations between databases were removed. The resulting variants were annotated by the Ensembl Variant Effect Predictor (VEP) (20). Those annotated as “inframe_insertion” or inframe_deletion” with biotype “protein coding” and a single protein consequence per variant were selected (n=3964; **Table 1, Table S1**). A subset of potentially novel variants from the DDD study, which were not present in either ClinVar or gnomAD (n=151), was used as an additional test set because these variants are unlikely to have been previously encountered by the tools.

**Figure 1.**
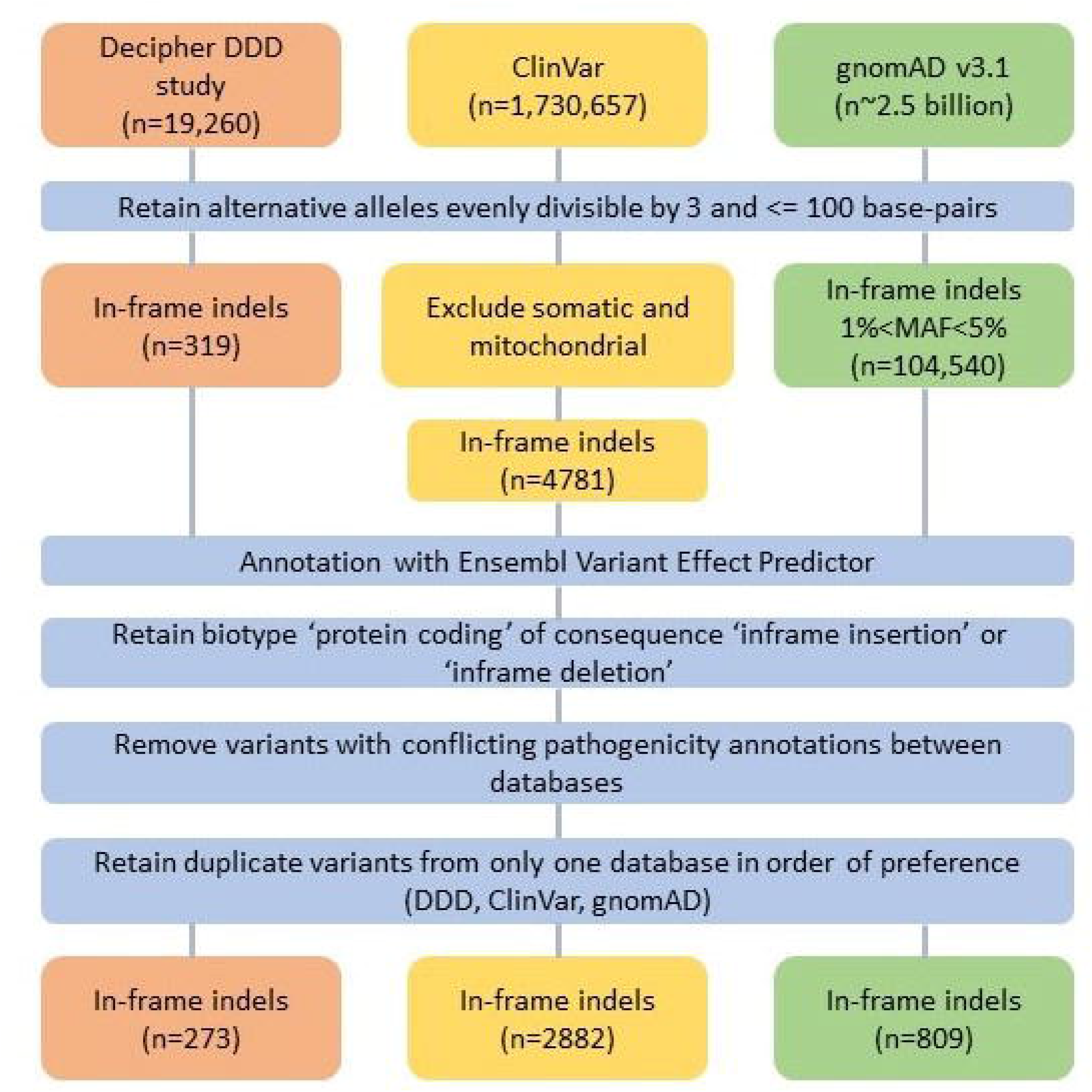
Flowchart of dataset construction. We included in-frame indels from ClinVar, gnomAD and the DDD study. SNV=single nucleotide variant, MAF=Minor allele frequency.

**Table 1.** Number of variants from each database included in our benchmark dataset.

| Database | Benign | Likely benign | Likely pathogenic | Pathogenic | Total |
| --- | --- | --- | --- | --- | --- |
| ClinVar | 424 | 803 | 796 | 859 | 2882 |
| DDD | 15 | 173 | 49 | 36 | 273 |
| gnomAD | 809 | 0 | 0 | 0 | 809 |
| **Total** | **1248** | **976** | **845** | **895** | **3964** |

### Tool selection and benchmarking

For inclusion in this study, pathogenicity prediction tools were identified from the literature and had to be either i) accessible through a webserver or ii) downloadable for use on a local server. We evaluated the performance of nine pathogenicity prediction tools, using their default classification threshold criteria: CADD (6), CAPICE (24), PROVEAN (25), FATHMM-indel (26), MutationTaster2 (27), MutPred-Indel (28), SIFT-indel (12), VEST-indel (29), and VVP (30) (**Table 2**). Standard performance metrics (sensitivity, specificity, positive and negative predictive values) and the Matthews Correlation Coefficient (MCC) (31) were calculated for all tools. Receiver-operator characteristics (ROC) and the area under the ROC curve (AUC) were determined for all tools apart from SIFT-indel and MutationTaster2 which produced binary classifications. All above analyses were repeated using the DDD-only subset. We also considered the effect of protein length on the ability of software to classify variants by grouping variants into four bins of amino acid length (1, 2-4, 5-10 and 11+).

**Table 2.** Pathogenicity predictors and default or recommended classification thresholds used in this study.

| Software | Input data format | Command line interface | Genome Build (GRCh) | Benign threshold | Pathogenic threshold |
| --- | --- | --- | --- | --- | --- |
| **CADD** | VCF | + | 37/38 | <20 | >=20 |
| **CAPICE** | VEP annotated TSV | + | 37/38 | <0.02 | >=0.02 |
| **FATHMM-indel** | VCF | - | 37 | >=0.5 | <0.5 |
| **MutPred-Indel** | Peptide (FASTA) | + | 37/38 | <0.672 | >=0.672 |
| **MutationTaster2** | VCF/genomic position/transcript specific | - | 37 | Benign | Deleterious |
| **PROVEAN** | VCF subset (CSV) | + | 37 | >2.5 | <=2.5 |
| **SIFT-indel** | VCF subset (CSV) | - | 37/38 | Neutral | Damaging |
| **VEST-indel** | VCF | + | 37/38 | <0.5 | >=0.5 |
| **VVP** | VCF | + | 37/38 | <57 | >=57 |

VCF=Variant call format; VEP=Variant effect predictor;
TSV=tab separated values; CSV=comma separated values.

## RESULTS

### Benchmark datasets contained a good balance of pathogenic and benign insertions and deletions

Our dataset consisted of 3964 small in-frame indels, including 1246 insertions and 2718 deletions from gnomAD (n=809), ClinVar (n=2882) and DDD (n=273) (**Figure 1**). Of these, 2224 were B/LB and 1740 were P/LP ranging in size from 1-48 amino acids for insertions and 1-66 amino acids for deletions (**Figure 2**). The longest pathogenic and benign deletions were 32 and 66 residues, and the longest pathogenic and benign insertions were 28 and 48 residues, respectively. Variants were distributed across 1820 protein-coding genes (mean=2.18, SD=3.64, min=1, max=66). The proportion of benign/pathogenic variants varied across genes linked with monogenic disease. Some genes had almost exclusively benign variants in our dataset, e.g. *DSPP* [MIM:125485] (B/LB=25, P/LP=0) and *ARID1B* [MIM:614556] (B/LB=62, P/LP=1); some had almost exclusively pathogenic variants, e.g. *LDLR* [MIM:606945] (P/LP=63, B/LB=0), *FBN1* [MIM: 134797] (P/LP=20, B/LB=0) and *NF1* [MIM:613113] (P/LP=17, B/LB=0); and some had similar numbers of pathogenic and benign variants, e.g. *CREBBP* [MIM:600140] (P/LP=10, B/LB=10) and *ARX* [MIM:300382] (P/LP=9, B/LB=10). The DDD-only dataset consisted of 151 novel in-frame indels, including 81 B/LB and 70 P/LP variants ranging in size from 1-11 and 1-13 amino acids for insertions and deletions, respectively, all in genes where rare deleterious variants are known to cause developmental disorders.

**Figure 2.**
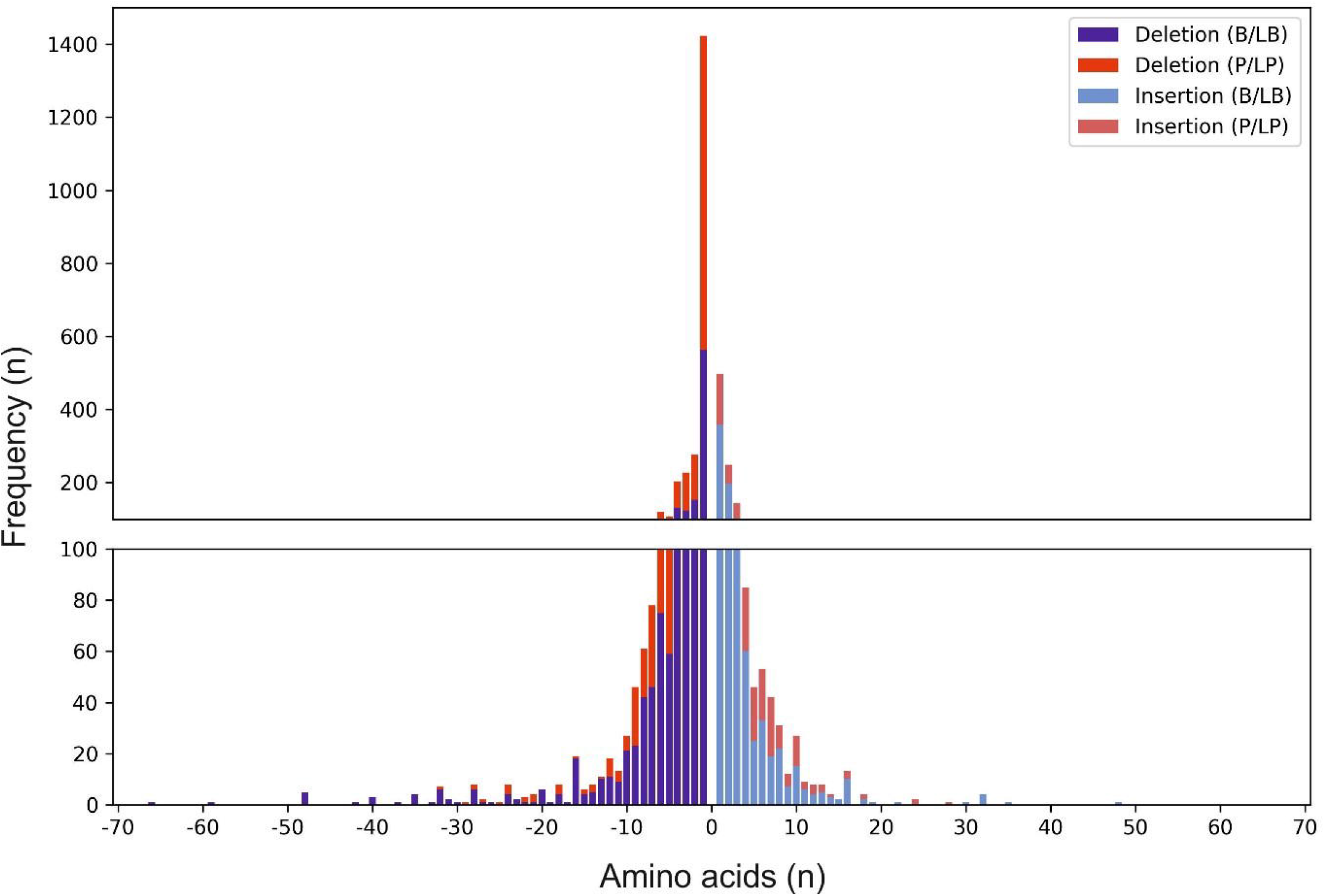
Histogram showing length and pathogenicity classification of our benchmark dataset. In-frame indels (n=3964, deletions=2718, insertions=1246) were taken from gnomAD, ClinVar and the DDD study. B/LB = benign/likely benign (blue); P/LP = pathogenic/likely pathogenic (red).

### Performance was generally high across all tools using our full dataset, but some tools performed substantially worse using a smaller, novel variant dataset

For the full dataset, 3615-3963 (91-99%) of variants were classified by each tool and 3522 (89%) were classified by every tool. Of the latter, 556 (15.8%) were universally categorised correctly by all nine tools as pathogenic (n=179, 5.1%) or benign (n=377, 10.7%). Sensitivity and specificity ranged from 0.30-0.99 and 0.61-0.97, respectively (**Table 3, Figure 3A**). For the smaller DDD-only novel dataset, 143-151 (95-100%) variants were classified by each tool and 141 (93%) were classified by every tool. Of these, 14 (9.9%) were universally categorised correctly by all nine tools as pathogenic (n=8, 5.7%) or benign (n=6, 4.2%). Sensitivity ranged from 0.24-0.97 and specificity range from 0.14-0.8 (**Table 3, Figure 3B**).

**Table 3.** Performance metrics for all indel pathogenicity prediction tools tested.

| Tool | TP | FP | TN | FN | Total (%) | Sensitivity | Specificity | LR+ | LR- | PPV | NPV | AUC | MCC |
| --- | --- | --- | --- | --- | --- | --- | --- | --- | --- | --- | --- | --- | --- |
| *All variants (1740 pathogenic; 2224 benign)* |  |  |  |  |  |  |  |  |  |  |  |  |  |
| **CADD** | 852 | 176 | 2047 | 886 | 3961 (99.9) | 0.49 | 0.92 | 6.19<sup>\$</sup> | 0.55 | 0.83 | 0.70 | 0.86 | 0.47 |
| **CAPICE** | 1611 | 715 | 1500 | 129 | 3955 (99.7) | 0.93 | 0.68 | 2.87<sup>+</sup> | 0.11<sup>+</sup> | 0.69 | 0.92 | 0.91 | 0.61 |
| **FATHMM-Indel** | 1626 | 572 | 1622 | 111 | 3931 (99.2) | 0.94 | 0.74 | 3.59<sup>+</sup> | 0.09<sup>+</sup> | 0.74 | 0.94 | 0.91 | 0.68 |
| **MutPred-Indel** | 516 | 72 | 2116 | 1211 | 3915 (98.8) | 0.30 | 0.97 | 9.08<sup>\$</sup> | 0.73 | 0.88 | 0.64 | 0.81 | 0.37 |
| **MutationTaster2** | 1675 | 99 | 1976 | 36 | 3786 (95.5) | 0.98 | 0.95 | 20.52<sup>\$</sup> | 0.02<sup>\$</sup> | 0.94 | 0.98 | - | 0.93 |
| **Provean** | 1482 | 581 | 1470 | 82 | 3615 (91.2) | 0.95 | 0.72 | 3.35<sup>+</sup> | 0.07<sup>\$</sup> | 0.72 | 0.95 | 0.93 | 0.66 |
| **SIFT-Indel** | 1406 | 827 | 1287 | 313 | 3833 (96.7) | 0.82 | 0.61 | 2.09 | 0.30<sup>+</sup> | 0.63 | 0.80 | - | 0.43 |
| **VEST-indel** | 1560 | 385 | 1758 | 179 | 3882 (98.0) | 0.90 | 0.82 | 4.99<sup>+</sup> | 0.13<sup>+</sup> | 0.80 | 0.91 | 0.93 | 0.71 |
| **VVP** | 1720 | 728 | 1481 | 20 | 3949 (99.6) | 0.99 | 0.67 | 3.00<sup>+</sup> | 0.02<sup>\$</sup> | 0.70 | 0.99 | 0.87 | 0.67 |

| **DDD subset (70 pathogenic; 81 benign)** |  |  |  |  |  |  |  |  |  |  |  |  |  |
| --- | --- | --- | --- | --- | --- | --- | --- | --- | --- | --- | --- | --- | --- |
| **CADD** | 45 | 16 | 65 | 25 | 151 (100) | 0.64 | 0.80 | 3.25<sup>+</sup> | 0.45 | 0.74 | 0.72 | 0.78 | 0.45 |
| **CAPICE** | 64 | 44 | 37 | 6 | 151 (100) | 0.91 | 0.46 | 1.68 | 0.19<sup>+</sup> | 0.59 | 0.86 | 0.82 | 0.41 |
| **FATHMM-indel** | 66 | 38 | 43 | 4 | 151 (100) | 0.94 | 0.53 | 2.01 | 0.11<sup>+</sup> | 0.63 | 0.91 | 0.74 | 0.51 |
| **MutPred-Indel** | 17 | 16 | 64 | 53 | 150 (99.3) | 0.24 | 0.80 | 1.21 | 0.95 | 0.52 | 0.55 | 0.64 | 0.05 |
| **MutationTaster2** | 50 | 19 | 61 | 19 | 149 (98.7) | 0.72 | 0.76 | 3.05<sup>+</sup> | 0.36<sup>+</sup> | 0.72 | 0.76 | - | 0.49 |
| **Provean** | 60 | 32 | 45 | 6 | 143 (94.7) | 0.91 | 0.58 | 2.19 | 0.16<sup>+</sup> | 0.65 | 0.88 | 0.86 | 0.51 |
| **SIFT-indel** | 60 | 39 | 41 | 10 | 150 (99.3) | 0.86 | 0.51 | 1.76 | 0.28<sup>+</sup> | 0.61 | 0.80 | - | 0.39 |
| **VEST-indel** | 62 | 29 | 51 | 8 | 150 (99.3) | 0.89 | 0.64 | 2.44<sup>+</sup> | 0.18<sup>+</sup> | 0.68 | 0.86 | 0.87 | 0.53 |
| **VVP** | 68 | 70 | 11 | 2 | 151 (100) | 0.97 | 0.14 | 1.12 | 0.21<sup>+</sup> | 0.49 | 0.85 | 0.64 | 0.19 |

Findings from the entire dataset are included in the top table, and just the novel (DDD-only) subset
in the bottom table. Relative strength of likelihood ratios for application of ‘strong’ or ‘moderate’
evidence under the ACMG/ACGS variant classification criteria(32) are denoted as: <sup>§</sup>High relative
strength, <sup>+</sup>Medium relative strength. **Figure S1** in **Additional File 1** shows ROC-AUC curves.
TP=True positive; FP=False positive; TN=True negative; FN=False negative; LR+=positive likelihood
ratio; LR-=negative likelihood ratio; PPV=Positive predictive value; NPV=negative predictive value;
AUC=Area under the curve; MCC=Matthews correlation coefficient.

**Figure 3.**
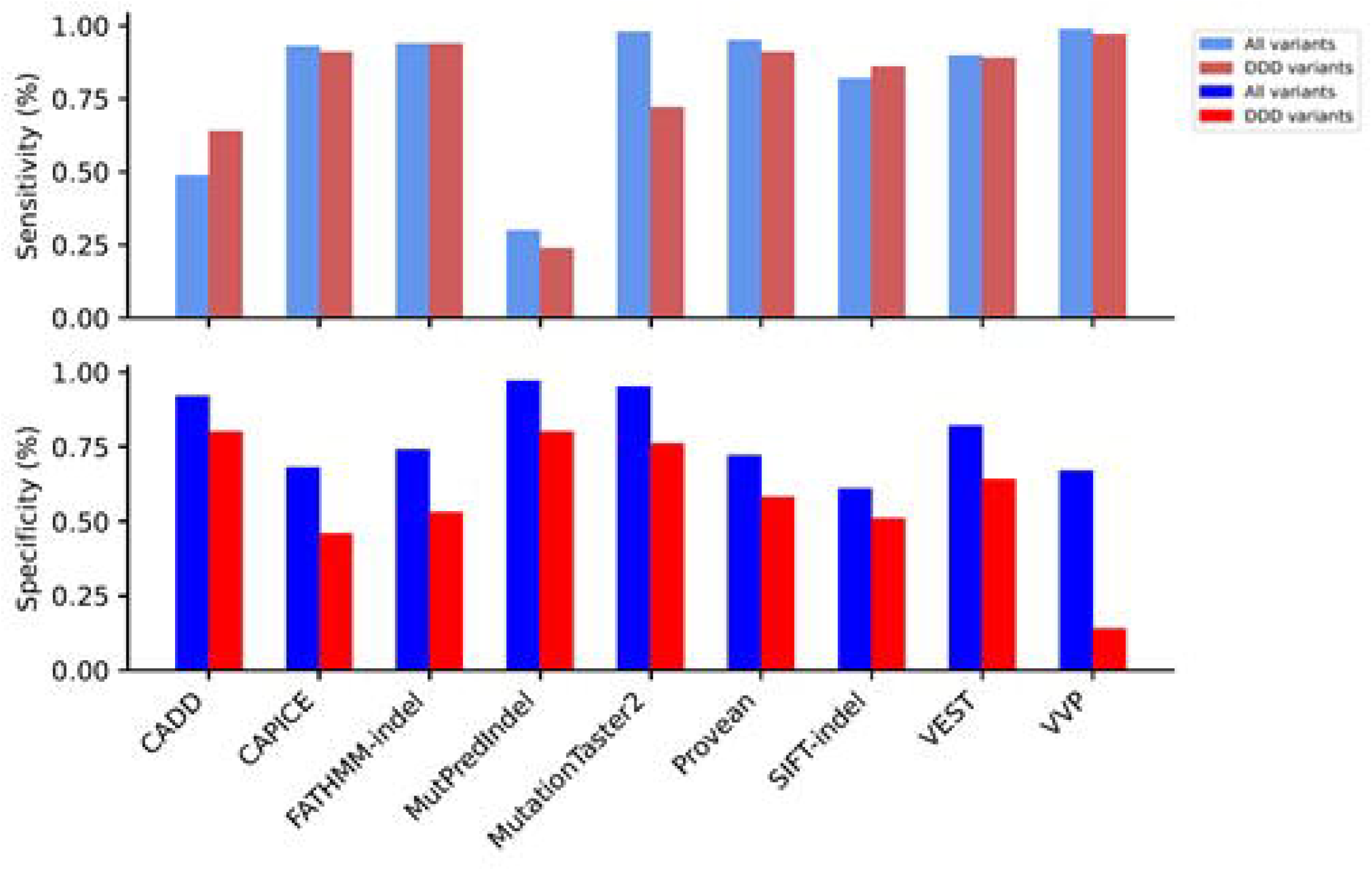
Performance of pathogenicity prediction tools for in-frame indels. **(A)** Sensitivity and **(B)** specificity of nine pathogenicity prediction tools based on classification of 3964 in-frame-indels from ClinVar, DDD and gnomAD databases (blue), as well as a DDD-only subset of 151 variants (red).

Sensitivity decreased for most tools between the full dataset and the DDD-only subset, apart from FATHMM-indel that remained the same (0.94), as well as CADD and SIFT-indel which increased from 0.49 to 0.64 and 0.82 to 0.86, respectively; MutationTaster2 showed the largest decrease in sensitivity from 0.98 to 0.72. Specificity decreased for all tools between the two datasets with CADD and SIFT-indel decreasing the least from 0.92 to 0.80 and 0.61 to 0.51, respectively; VVP decreased the most from 0.67 to 0.14. These observations were recapitulated in the MCC metric, where VVP and MutationTaster2 decreased the most by 0.48 and 0.44, whereas CADD and SIFT-Indel decreased the least by 0.02 and 0.04, respectively.

### Tool performance was generally independent of indel length

We investigated the tools’ performance for insertions and deletions separately, and whether their performance was influenced by indel length (grouped into bins of 1, 2-4, 5-10 and 11+ amino acids inserted/deleted). We observed very little difference in performance between groups of variants (**Figure S2** and **S3** in **Additional File 1** shows this in more detail), despite an increase in the proportion of benign variants with increasing indel length.

## DISCUSSION

We tested the performance of nine pathogenicity prediction tools on a dataset of 3964 in-frame indels and a smaller subset of 151 novel, clinically classified indels that are not readily accessible from public databases. We show that the performance of these tools is generally good across a range of indel lengths, with AUCs of 0.81-0.93. As expected, most tools performed less well in the smaller novel subset, with AUCs of 0.64-0.87, which likely reflects the use of publicly accessible datasets in the tools’ classification method or training data.

Of the nine tools tested, FATHMM-indel, CAPICE, VEST-indel and PROVEAN performed comparably well, although PROVEAN and VEST-indel classified fewer variants than CAPICE and FATHMM-indel. It should be noted that some tools (e.g. CADD, CAPICE, MutationTaster2, PROVEAN, VVP) were not designed specifically for use with in-frame indels and were likely trained primarily on SNVs, whilst other tools (e.g. VEST-indel, FATHMM-indel, MutPred-Indel, SIFT-indel) were optimised particularly for the classification of indels. We have previously demonstrated that standard pathogenicity predictors such as SIFT and Polyphen-2 classified missense variants with AUCs of between 0.85-0.87 for a publicly accessible “open” dataset, and 0.70-0.72 for a restricted access “clinical” dataset (33), which is a comparable performance to the indel pathogenicity predictors tested here. However, the newer meta-predictors Revel (34)and ClinPred (35) produced AUCs of 0.97-0.99 and 0.82-0.81 for open and clinical datasets of missense variants (33), respectively, outperforming all the indel pathogenicity prediction tools tested here. Nonetheless, similar to many missense pathogenicity predictors, the likelihood ratios calculated for in-frame indel predictors using our dataset (**Table 3**) support their use at either ‘supporting’ or ‘moderate’ towards the PP3 and BP4 criteria of the ACMG/ACGS recommendations, although none of the tools reach the moderate threshold in the DDD subset (32, 36).

We found that the pathogenicity predictor tools varied substantially in input requirements and their ease of use. For example, seven of the tools tested require variants to be uploaded in VCF format as input and five of these also offer a downloadable command line interface (**Table 2**). Tools with these two features are typically well suited for integration into analysis pipelines; however, ease of installation, additional required software and metadata dependencies varied. For example, MutPred-Indel contained all the necessary metadata to make pathogenicity predictions, but required variants to be input in FASTA format, which is not routinely used in a clinical genetic testing setting, as well as installation of a specific version of MATLAB. Similarly, PROVEAN offered a command line option but also required local installations of NCBI-Blast, and the NCBI nr protein database. The requirement for advanced bioinformatics skills to operate a tool will adversely affect its utility, particularly for routine diagnostics use. In contrast, CAPICE used an Ensembl-VEP annotated TSV file as input, and we found it the easiest to install and quickest to use.

Like other comparable studies, we were limited by several factors. Firstly, the veracity of the variant classifications taken from ClinVar, the DDD study and gnomAD is uncertain, and our benchmark dataset may include some erroneous variant classifications. We tried to minimise this issue by incorporating data from three different databases and by using minor allele frequency thresholds for benign variants. Secondly, unlike missense variants caused by SNVs, in-frame indels are comparatively rare and are harder to detect robustly using NGS, and thus our dataset is relatively small. Evaluation of the performance of the tools versus indel length was further limited by the inverse correlation between frequency and variant length in the dataset, which limits the interpretability of tool performance for larger indels. Although large (>100 base-pair) in-frame indels exist, and may be either both benign or pathogenic, these are difficult to detect using short-read NGS technologies, so were largely absent from the databases used here. Finally, not all variants in our dataset were in monogenic disease genes, particularly those from gnomAD, which potentially introduces a bias for tools that use gene-level data for classification. However, around 75% of genes present in our dataset contained variants from at least two of the databases, and a sensitivity analysis using only these variants produced similar results (data not shown).

## CONCLUSIONS

We have shown that numerous *in silico* pathogenicity prediction tools perform well for in-frame indels using a benchmark dataset. Our findings are consistent with previous studies (24, 26, 29) and, to the best of our knowledge, represent the largest independent assessment to date of pathogenicity predictors for in-frame indels. We suggest that genomic diagnostic laboratories should consider incorporating these tools, in the same manner as missense prediction tools, to aid variant classification.

## Data Availability

The publically available variant datasets are available from gnomAD, ClinVar. Genomic datasets from the DDD Study are available under managed access for research into developmental disorders via the European Genome-phenome Archive (EGAS00001000775). Individual pathogenic/likely pathogenic variants are openly accessible with phenotypes via DECIPHER.

https://gnomad.broadinstitute.org/

https://www.ncbi.nlm.nih.gov/clinvar/

https://www.deciphergenomics.org/

## LIST OF ABBREVIATIONS

ACGS: UK Association for Clinical Genomic Science
ACMG: American College of Medical Genetics and Genomics
AMP: Association for Molecular Pathology
AUC: Area Under the Curve
B: Benign
CSV: Comma Separated Values
DDD: Deciphering Developmental Disorders
FP: False Positive
FN: False Negative
LB: Likely Benign
LP: Likely Pathogenic
LR+: Positive Likelihood Ratio
LR: Negative Likelihood Ratio
MCC: Matthews Correlation Coefficient
MIM: Mendelian Inheritance in Man
NGS: Next Generation Sequencing
NPV: Negative Predictive Value
P: Pathogenic
PPV: Positive Predictive Value
ROC: Receiver Operator curve
SD: Standard Deviation
SNV: Single Nucleotide Variant
TP: True Positive
TN: True Negative
TSV: Tab Separated Values
VCF: Variant Call Format
VEP: Variant Effect Predictor

## DECLARATIONS

### ETHICS APPROVAL AND CONSENT TO PARTICIPATE

The study has UK Research Ethics Committee approval (10/H0305/83, granted by the Cambridge South REC, and GEN/284/12 granted by the Republic of Ireland REC) and all participants provided consent.

### CONSENT FOR PUBLICATION

Not applicable

### AVAILABILITY OF DATA AND MATERIALS

The publically available variant datasets are available from gnomAD (22), ClinVar (10). Genomic datasets from the DDD Study are available under managed access for research into developmental disorders via the European Genome-phenome Archive (EGAS00001000775). Individual pathogenic/likely pathogenic variants are openly accessible with phenotypes via DECIPHER (37). All variants in the final dataset are included in Additional File 2.

### AUTHOR’S CONTRIBUTIONS

CFW conceived and designed the study and provided the datasets. MW performed an initial analysis and write-up using a subset of tools, with assistance from ACG. SC performed the full data analysis and wrote the first draft of the manuscript. SC and CFW finalised the manuscript, and all authors approved it.

## ACKNOWLEDGEMENTS

We wish to thank all the patients and family members whose data were used in the study. The DDD study presents independent research commissioned by the Health Innovation Challenge Fund (grant number HICF-1009-003), a parallel funding partnership between the Wellcome Trust and the Department of Health, and the Wellcome Trust Sanger Institute (grant no. WT098051). See Nature 2015;519:223–8 or www.ddduk.org/access.html for full acknowledgement.

## ADDITIONAL FILES

### Additional File 1

#### File name and format

“Additional File 1.docx”

#### Title and description of data

Figure S1 - ROC curves and AUCs using (A) 3964 in-frame indels from gnomAD, ClinVar and the DDD study (B) 151 in-frame indels only observed in the DDD study. Only tools producing continuous data as output are plotted (n=7).

Figure S2. Performance of nine pathogenicity prediction tools versus increasing variant length in a dataset of 3964 in-frame indels aggregated from gnomAD, ClinVar and the DDD study.

Figure S3. Sensitivity and specificity of 9 pathogenicity prediction algorithms with increasing variant length in a dataset of in-frame (A) deletions (n=2718) and (B) insertions (n=1246).

## ADDITIONAL FILE 2

### File name and format

“Additional File 2.xlsx”

#### Title and description of data

Inframe-indels and pathogenicity predictions from tools used in this study.

